# Incremental value of bilateral ankle-brachial index for ten-year heart and cerebrovascular mortality prediction: an exploratory temporal benchmark in NHANES

**DOI:** 10.64898/2026.10.30.26366960

**Authors:** Lingsen You, Yujun Guo, Xinyu Zhong, Zisu Peng, Wentong Wang, Li Shen, Junbo Ge

## Abstract

**Background:** Ankle-brachial index (ABI) is associated with systemic vascular disease, but its incremental predictive value depends on the clinical comparator, endpoint and validation design.

**Objective:** To test whether a bilateral ABI feature block improves a nonlaboratory clinical benchmark for ten-year death from diseases of the heart or cerebrovascular disease, and whether a limited nonlinear learner improves the same chronological evaluation.

**Methods:** We rebuilt public NHANES 1999–2004 examination data linked to the 2019 public-use mortality file. Adults aged at least 40 years with bilateral ABI values in [0.20, 2.50] and ascertainable 120-month status formed a survey-weighted analytic subset. Models were trained on 1999–2002, tuned within development cycles, and evaluated on 2003–2004. Primary models were linear clinical logistic regression, flexible clinical logistic regression, flexible clinical plus ABI regression, and histogram gradient boosting plus ABI. Point estimates used full evaluation weights; conditional intervals used 32-replicate Fay balanced repeated replication with 15 design degrees of freedom. The study was not prospectively registered.

**Results:** The corrected cohort contained 6,807 participants (4,617 development; 2,190 temporal evaluation) and 409 primary events, including 139 in evaluation. Evaluation AUROC was 0.834 for flexible clinical regression and 0.834 after ABI addition. The paired ABI increment was -0.0007 (95% CI -0.0047 to 0.0032); the paired Brier difference was 0.0008 (95% CI -0.0003 to 0.0018). Boosting plus ABI had AUROC 0.822; versus regression with the same predictor block, its AUROC difference was -0.0117 (95% CI -0.0379 to 0.0144). Its Brier difference was -0.000661 (95% CI -0.001322 to -7.53e-08); the upper limit was extremely close to zero. Neither primary hypothesis of improvement in both metrics was supported.

**Conclusions:** In this corrective, exploratory chronological benchmark, bilateral ABI did not improve discrimination or Brier score beyond a matched flexible clinical model. The result does not negate ABI associations, establish equivalence, or support treatment decisions. It supports transparent evaluation of incremental information in a new cohort rather than claims of clinical utility.

## Introduction

Peripheral, coronary and cerebrovascular disease share systemic risk, which motivates a panvascular view of interventional medicine. Recent work from the Zhongshan group places device, vascular and organ-level questions within a broader panvascular programme [1–3]. Those papers are clinical or conceptual context; none is evidence for the present NHANES model. ABI is attractive for risk stratification because it is inexpensive and captures lower-extremity arterial physiology. Numerous cohorts and a participant-level meta-analysis associate abnormal ABI with mortality and cardiovascular events [4–11].

The incremental prediction question is narrower than the association question. Jazayeri and colleagues already evaluated ABI risk stratification in NHANES, including continuous net reclassification [5], and a recent NHANES machine-learning study also investigated mortality prediction [12]. Therefore, the contribution claimed here is a reproducible comparison of a defined bilateral ABI block against matched nonlaboratory clinical models, with discrimination, proper scoring, calibration, decision summaries and design-aware paired uncertainty. The study is an exploratory corrective reanalysis after earlier draft outcomes had been inspected; it is not a preregistration or confirmatory independent validation.

We asked two testable questions. H1 was that adding bilateral ABI summaries to a flexible clinical model would improve temporal-evaluation AUROC and Brier score. H2 was that a limited gradient-boosting model would improve both metrics over flexible regression using the same clinical-plus-ABI predictor block. These questions guided the corrective run, but remain exploratory because previous evaluation outcomes were known.

## Methods

### Design and data

This secondary analysis used de-identified NHANES 1999–2004 examination files and the NCHS 2019 public-use linked mortality file [19,20]. The files are publicly downloadable; no restricted records, direct personal identifiers or participant contact were used; public SEQN identifiers were used solely for file linkage. NHANES examination weights were pooled as two-thirds of WTMEC4YR for 1999–2002 and one-third of WTMEC2YR for 2003–2004. The analytic unit is a survey-weighted ABI-observed subset, not all US adults aged at least 40 years. Study size was determined by all eligible participants in the available files, with cycles assigned chronologically; no formal sample-size or power calculation was performed. The 270 development and 139 evaluation events limit precision and model stability, so the study cannot establish equivalence or exclude small increments. The 2019 public-use mortality release follows participants through 31 December 2019; the primary outcome here was truncated at 120 months after examination [20].

### Cohort and outcomes

We included participants aged at least 40 years with positive examination weight, linkage eligibility, known mortality status and ascertainable follow-up. Both ABI legs had to be observed and lie within 0.20–2.50. One 2001–2002 participant with a death within ten years but unavailable underlying cause was excluded from endpoint ascertainment. The sequential flow is shown in Figure 1. Development comprised 1999– 2002; 2003–2004 was the chronological evaluation set. The primary endpoint was cumulative death by 120 months from diseases of heart (underlying-cause code 001) or cerebrovascular disease (005). Death from other causes by 120 months was retained as a competing non-event; this estimand is not a cause-specific hazard. Survivors required at least 120 months of follow-up. All-cause death and five-year endpoints were exploratory sensitivities. Public-use mortality linkage can perturb selected follow-up times and causes for disclosure protection.

**Figure 1.**
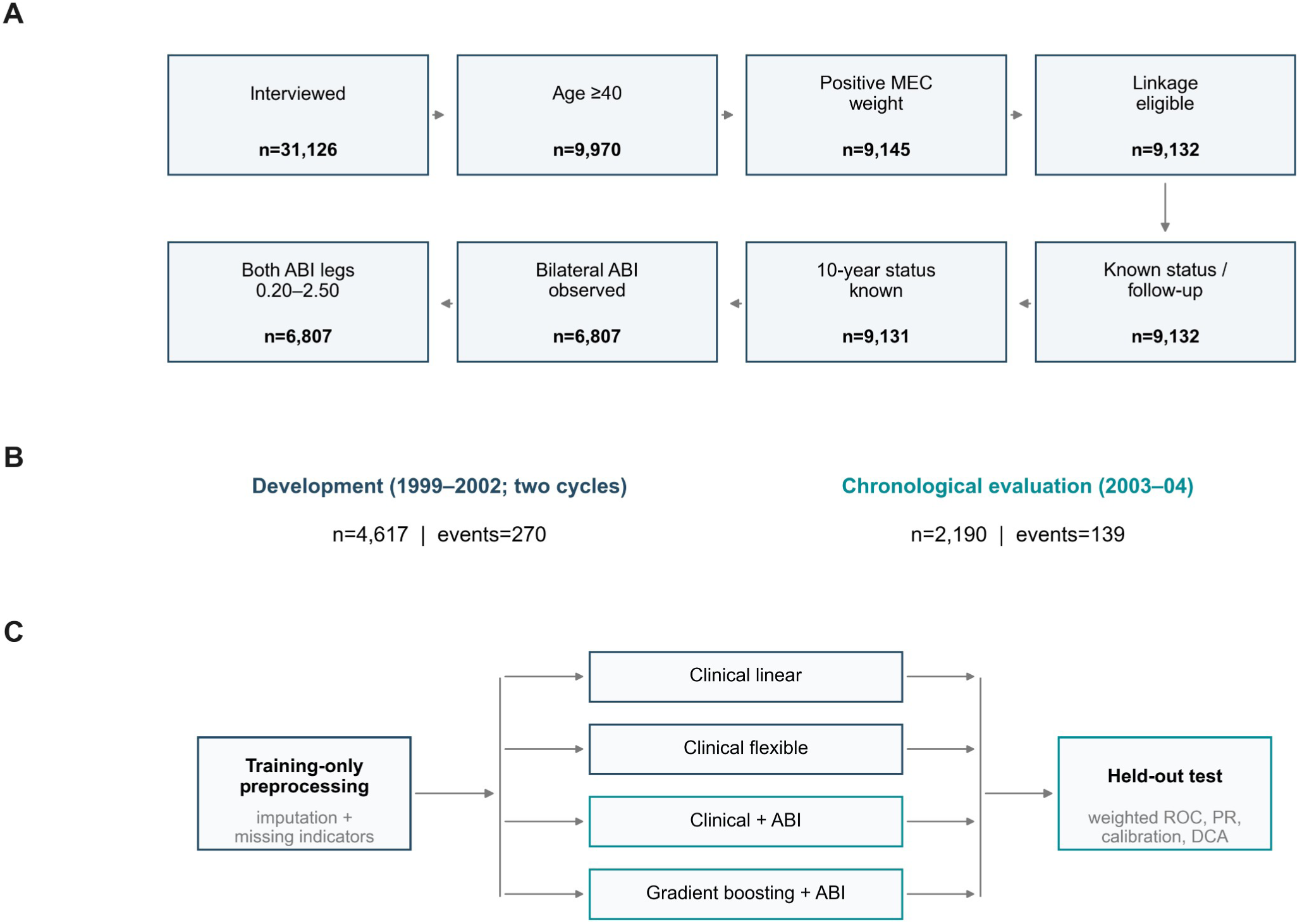
Study cohort, modeling workflow, and evaluation. (A) Sequential all-cycle analytic flow with retained counts after each source criterion. (B) Development sample (n=4,617; 270 events) and chronological evaluation sample from 2003–04 (n=2,190; 139 events). (C) Training-only preprocessing, clinical linear and flexible models, ABI extension, and held-out weighted evaluation.

### Predictors and models

The nonlaboratory clinical block comprised age, sex, race/ethnicity, education, poverty-income ratio, body mass index, waist circumference, mean systolic and diastolic pressure (all positive available readings, including the first), pulse, reported diabetes, hypertension, prior cardiovascular disease and smoking category. Smoking used the questionnaire skip rule: SMQ020=2 was never smoking regardless of structurally skipped SMQ040; other categories used SMQ040. Nominal race, education and smoking categories were one-hot encoded. ABI predictors were minimum ABI, absolute between-leg ABI difference and an indicator that either leg was at least 1.40. The high-ABI tail was descriptive because only six evaluation participants had minimum ABI at least 1.40.

We compared linear clinical logistic regression, flexible clinical logistic regression with cubic splines for age, systolic pressure and body mass index, flexible clinical plus ABI regression, and histogram gradient boosting plus ABI. C or boosting regularization was selected by weighted Brier score using 1999–2000 fitting and 2001–2002 tuning, then refit on all development participants. Imputation, missingness indicators, category encoding, scaling and spline knots were fitted inside the development pipeline only. Training weights were normalized to mean one for penalized fitting. Regression used ridge penalization with C in {0.01, 0.1, 1, 10}; all three main logistic models selected C=0.1. Cubic splines used four training-quantile knots and linear extrapolation. Boosting used 150 iterations, learning rate 0.04, minimum leaf size 35, no early stopping, and the development grid of 7 or 15 leaves crossed with L2=1 or 10; the selected configuration was 7 leaves and L2=10. Python and scikit-learn versions and all selected parameters are recorded in the execution manifest [21]. No mortality, follow-up, cause-of-death, cycle, PSU, stratum or participant identifier entered a feature matrix. After the primary results, we added a matched clinical-only boosting comparator and a strict high-ABI threshold sensitivity (>1.40); these post hoc additions used the same development-only tuning scheme and did not alter the primary models. Code and retained outputs accompany the manuscript; the word evaluation denotes the chronological test cycle, not an independently untouched external cohort.

### Evaluation and uncertainty

Primary metrics were weighted AUROC, average precision, Brier score [17], calibration-in-the-large (prediction-logit slope fixed at one) and calibration slope [16]. Full-weight evaluation values are the point estimates. Conditional evaluation uncertainty used Fay BRR with rho=0.5, 32 distinct Hadamard sign patterns, 15 two-PSU strata and t(15) intervals. The same replicate weights were used for paired model differences and domain analyses retained the full design map. These intervals condition on fitted models and omit development/model-selection uncertainty. Decision curves [15] are hypothetical net-benefit summaries; no intervention threshold was prespecified. Reporting was guided by TRIPOD+AI [13] and the risk-of-bias issues in PROBAST+AI [14]. An item-level reporting checklist with remaining limitations is included in the reproducibility archive; full guideline compliance is not claimed.

### Corrective revision and AI assistance

No prospective registration was performed; the corrective protocol and later amendment are provided in the source archive and postdate initial inspection of evaluation outcomes. No patient or public involvement is documented for the design, conduct, interpretation or reporting of this computational secondary analysis.

The earlier draft used generic missing-value recoding, incorrect pooled weights, incomplete smoking skip handling and an inconsistent resampling scheme. Those files and results are retained as superseded audit artifacts and are not used here. The original working plan emphasized five-year all-cause survival, whereas the earlier executed draft used ten-year heart/cerebrovascular mortality. This change is disclosed as exploratory. The revised protocol was recorded before the corrected run, but the evaluation outcomes had already been inspected. Codex assisted with analysis-code assembly, figure generation, prose editing and consistency checks; this assistance is reported separately from human author responsibility. The AI assistance included proposing the study question and analytic revision, developing and running code, drafting text, preparing numerical graphics and organizing reference checks. No synthetic participant records or simulated empirical outcomes were used. Internal agents performed computational and documentary checks only and are not peer reviewers or manuscript authors.

## Results

### Cohort and vascular phenotype

The rebuilt flow included 31,126 interviewed participants, 9,131 otherwise eligible participants before ABI exclusions and 6,807 participants in the analytic set. The development set contained 4,617 participants and 270 primary events; the evaluation set contained 2,190 participants and 139 events. Eighty-three age-77 participants and 41 participants with waist values of 77 or 99 cm were retained. A total of 2,324 otherwise eligible participants lacked acceptable bilateral ABI. Excluded participants were older and had more measured diabetes, hypertension and prior cardiovascular disease; the resulting risks should therefore be interpreted within the ABI-observed subset. Figure 2 presents phenotype support and development-to-evaluation case mix; the inclusion-selection comparison is provided in the supplement.

**Figure 2.**
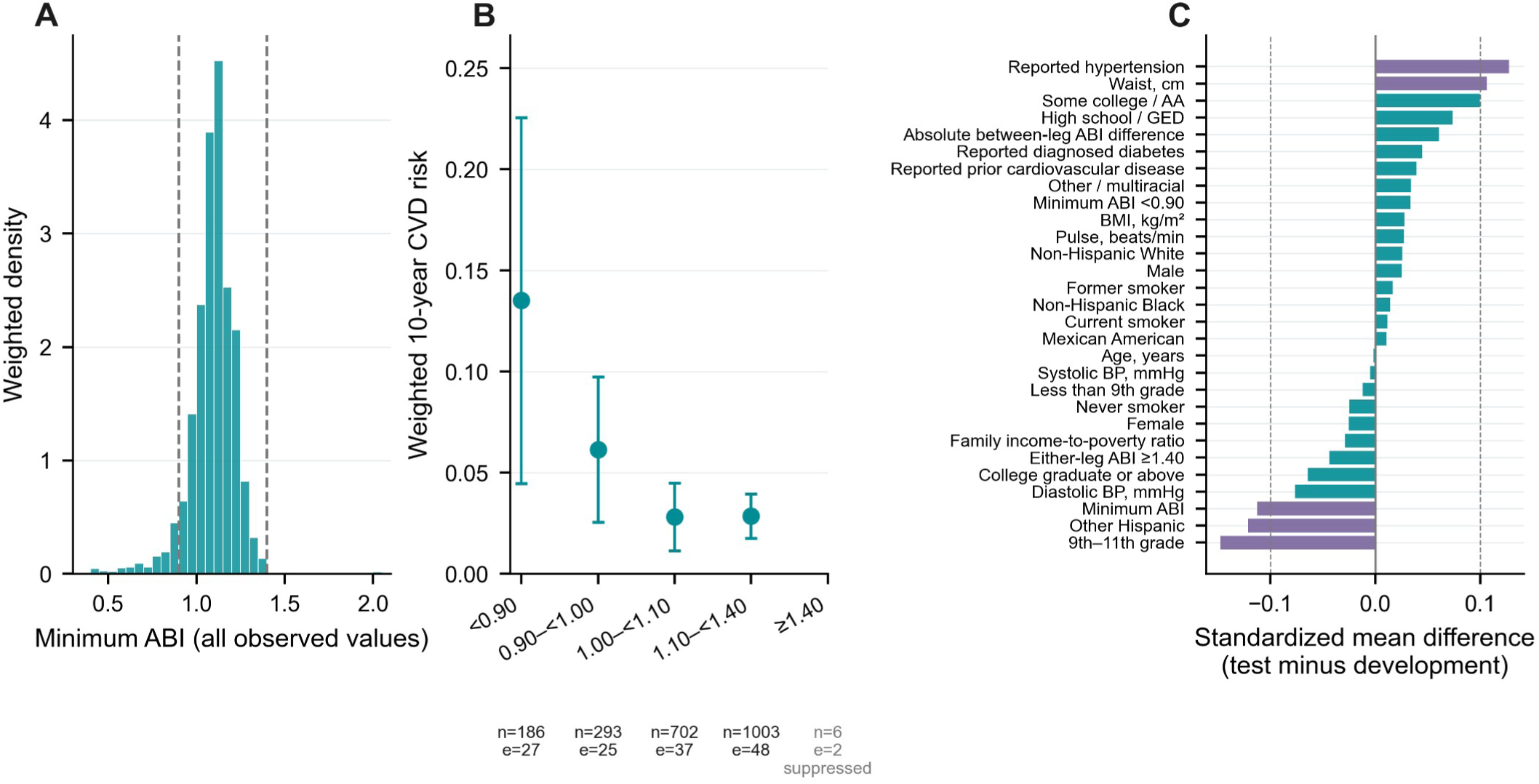
ABI distribution, risk by ABI category, and development-to-test case mix. (A) Survey-weighted density across the full observed minimum ABI range (0.33–2.07) among participants with measured ABI; dashed lines mark 0.90 and 1.40. (B) Weighted 10-year CVD risk and 95% intervals by ABI category, with unweighted category counts and events. Risk is suppressed for the ≥1.40 tail (n=6; 2 events) under the sparse-cell rule. (C) Standardized mean differences (test minus development); dashed vertical lines mark ±0.10.

### Primary temporal evaluation

The flexible clinical model had AUROC 0.834 (95% CI 0.786–0.882) and Brier score 0.0329. Adding ABI yielded AUROC 0.834 (95% CI 0.788–0.879) and Brier score 0.0337. The paired AUROC difference was -0.0007 (95% CI -0.0047 to 0.0032), and the paired Brier difference was 0.0008 (95% CI -0.0003 to 0.0018). Thus H1 was not supported by this evaluation. The linear clinical model had AUROC 0.830. Gradient boosting plus ABI had AUROC 0.822; versus regression with the same clinical-plus-ABI block, its paired AUROC difference was -0.0117 (95% CI -0.0379 to 0.0144). Its Brier score was lower by 0.000661 (difference 95% CI -0.001322 to -7.53e-08), with the upper bound extremely close to zero. H2, requiring improvement in both metrics, was not supported. Versus the flexible clinical-only model, the boosting AUROC difference was -0.0125 (95% CI -0.0407 to 0.0158). Figure 3 shows weighted ROC and precision–recall curves and paired contrasts.

**Figure 3.**
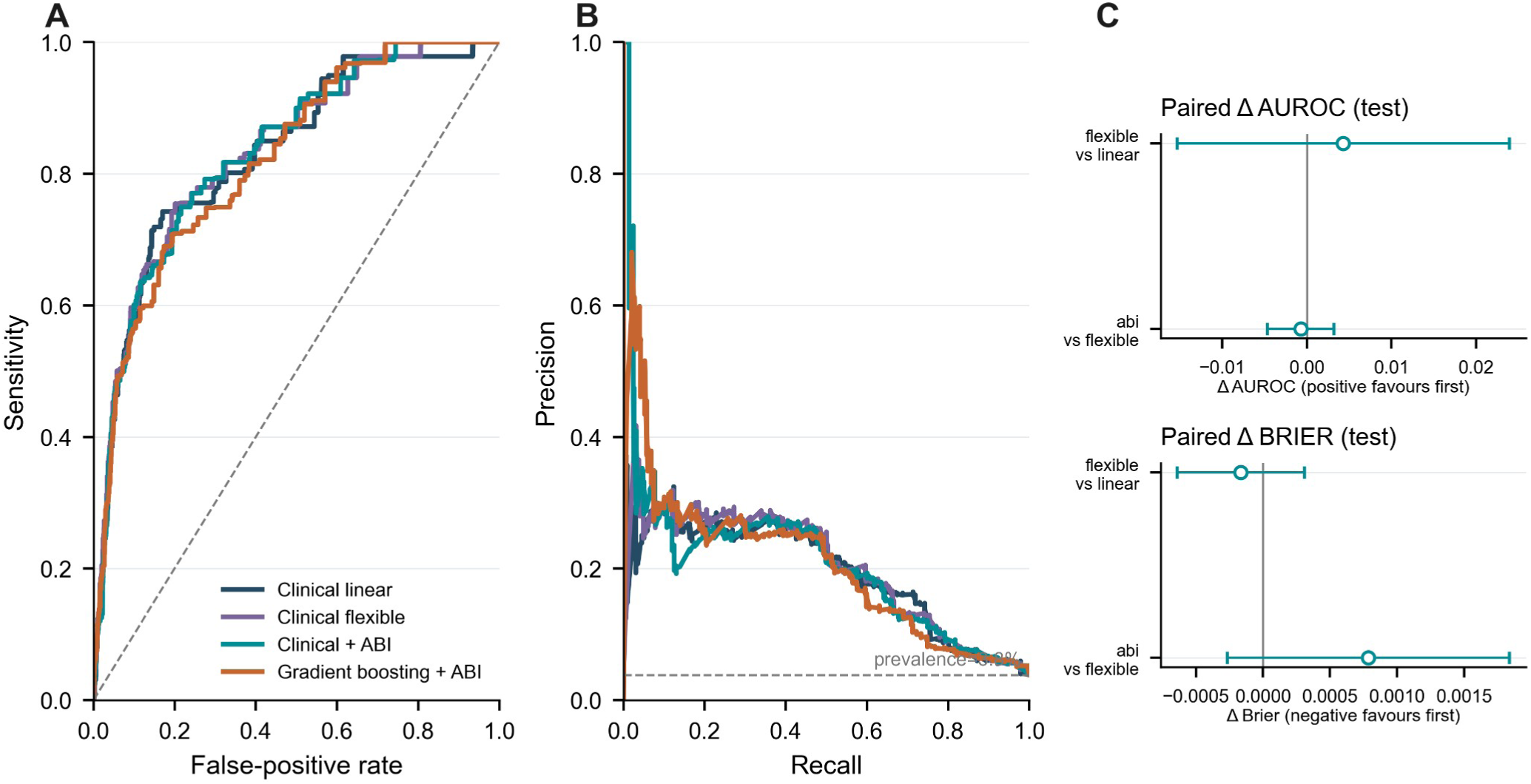
Discrimination and paired model contrasts in the chronological test sample. (A) Weighted receiver operating characteristic curves. (B) Weighted precision–recall curves; dashed reference is survey-weighted event prevalence (3.80%). (C) Paired differences and 95% intervals for AUROC and Brier score. Positive AUROC differences and negative Brier differences favor the first named model.

Calibration slopes were 0.90 for flexible clinical regression, 0.82 for clinical plus ABI, and 0.82 for boosting. Figure 4 shows calibration in the observed low-risk support rather than compressing the curves into a 0–1 axis. Decision curves in Figure 5 are descriptive; no threshold was selected and no deaths prevented or treatments assigned can be inferred.

**Figure 4.**
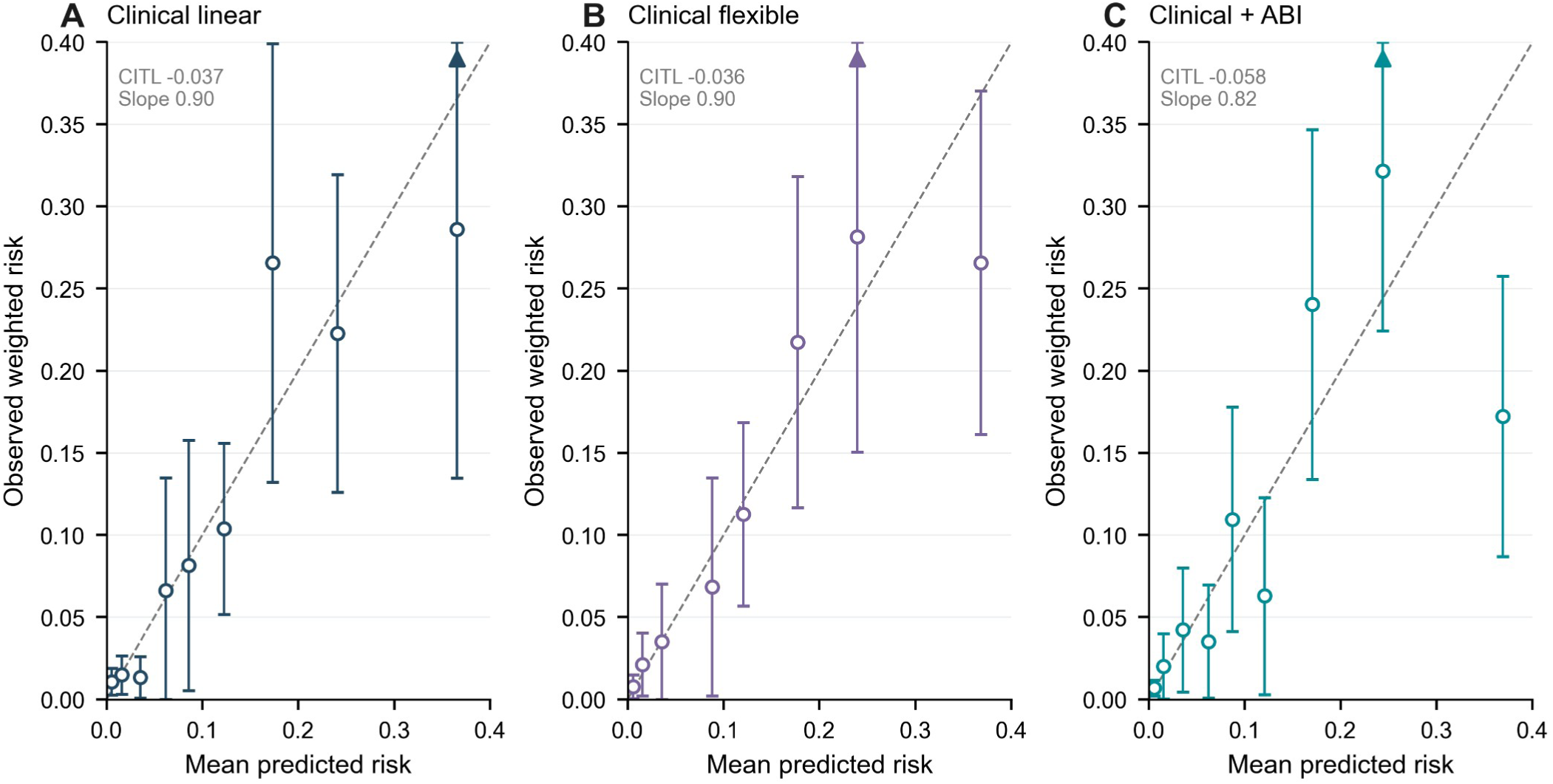
Calibration of three clinical regression models in the chronological test sample. (A) Clinical linear. (B) Clinical flexible. (C) Clinical + ABI. Points show observed weighted risk by supported prediction bin with 95% intervals; dashed line indicates perfect calibration. Upward triangles identify intervals with an upper confidence limit above the displayed 0.40 range. Insets report calibration-in-the-large and calibration slope. Bins with fewer than five events are suppressed; Supplementary Figure S4 shows the full scale.

**Figure 5.**
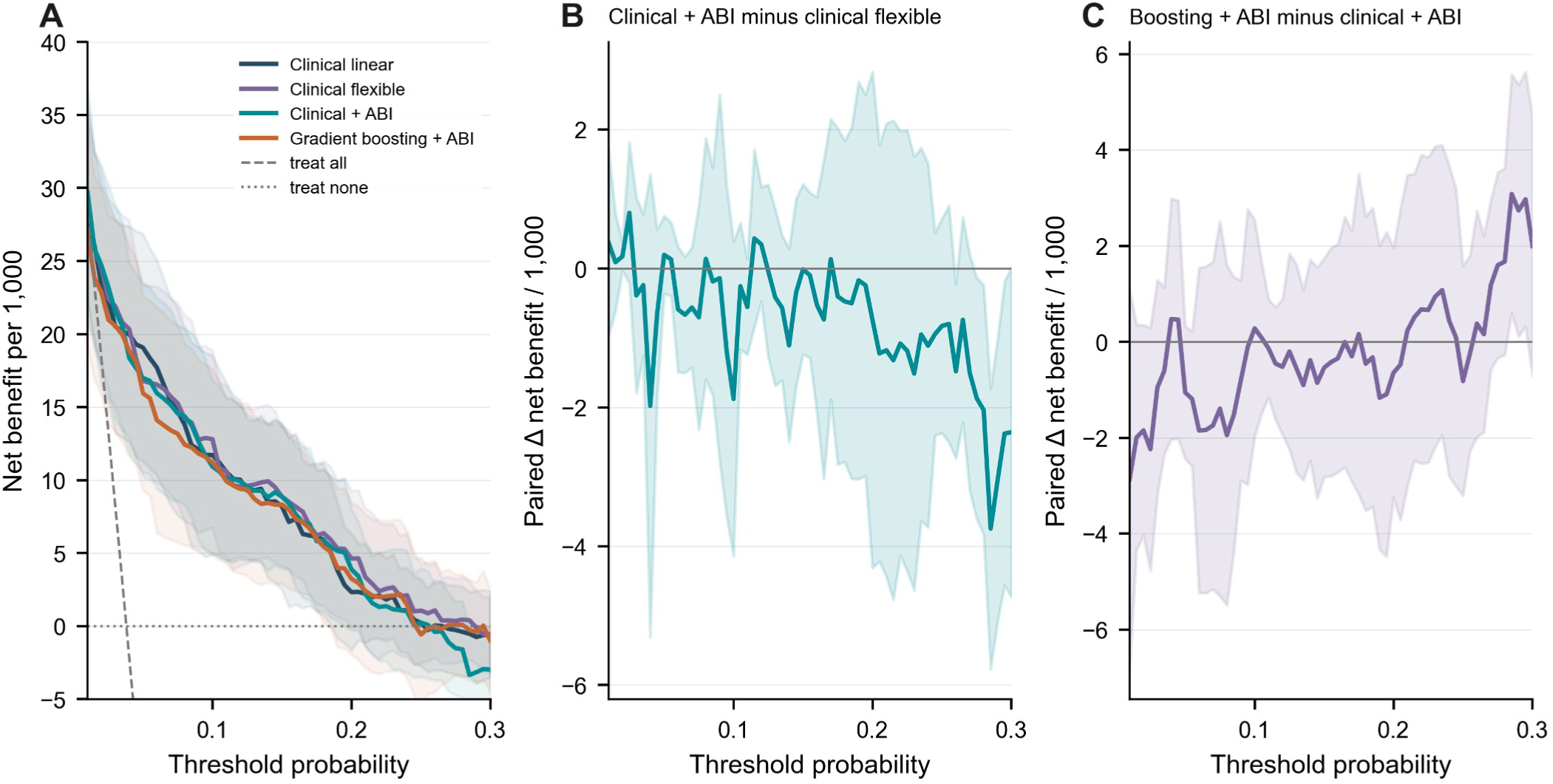
Decision-curve analysis across hypothetical threshold probabilities. (A) Absolute net benefit per 1,000 for each model, treat-all, and treat-none, displayed from −5 to 40 per 1,000; treat-all falls below the displayed range at high thresholds. (B) Paired net-benefit difference: clinical + ABI minus clinical flexible. (C) Gradient boosting + ABI minus clinical + ABI. Shaded regions show conditional 95% Fay BRR intervals. These are descriptive decision summaries, not treatment effects.

### Ablation, heterogeneity and sensitivity

Replacing the full ABI block with minimum ABI alone produced an AUROC difference of approximately −0.0006 versus the flexible clinical model; adding the bilateral range did not provide a stable improvement. Grouped ABI permutation preserves relationships among ABI-derived variables and is interpreted as predictive reliance, not biological importance (Figure 6). Exploratory subgroup and endpoint contrasts are shown in Figure 7; calibration, permutation reliance and missing-data sensitivities are detailed in Supplementary Figures S4, S5 and S6, respectively. The ten-year all-cause analysis produced a lower Brier score for clinical plus ABI than the flexible clinical model (difference -0.0011, 95% CI -0.0020 to -0.0002); this endpoint-specific signal was not used to replace the primary cause-specific conclusion. Five-year and complete-case contrasts were imprecise and did not establish a consistent ABI increment.

**Figure 6.**
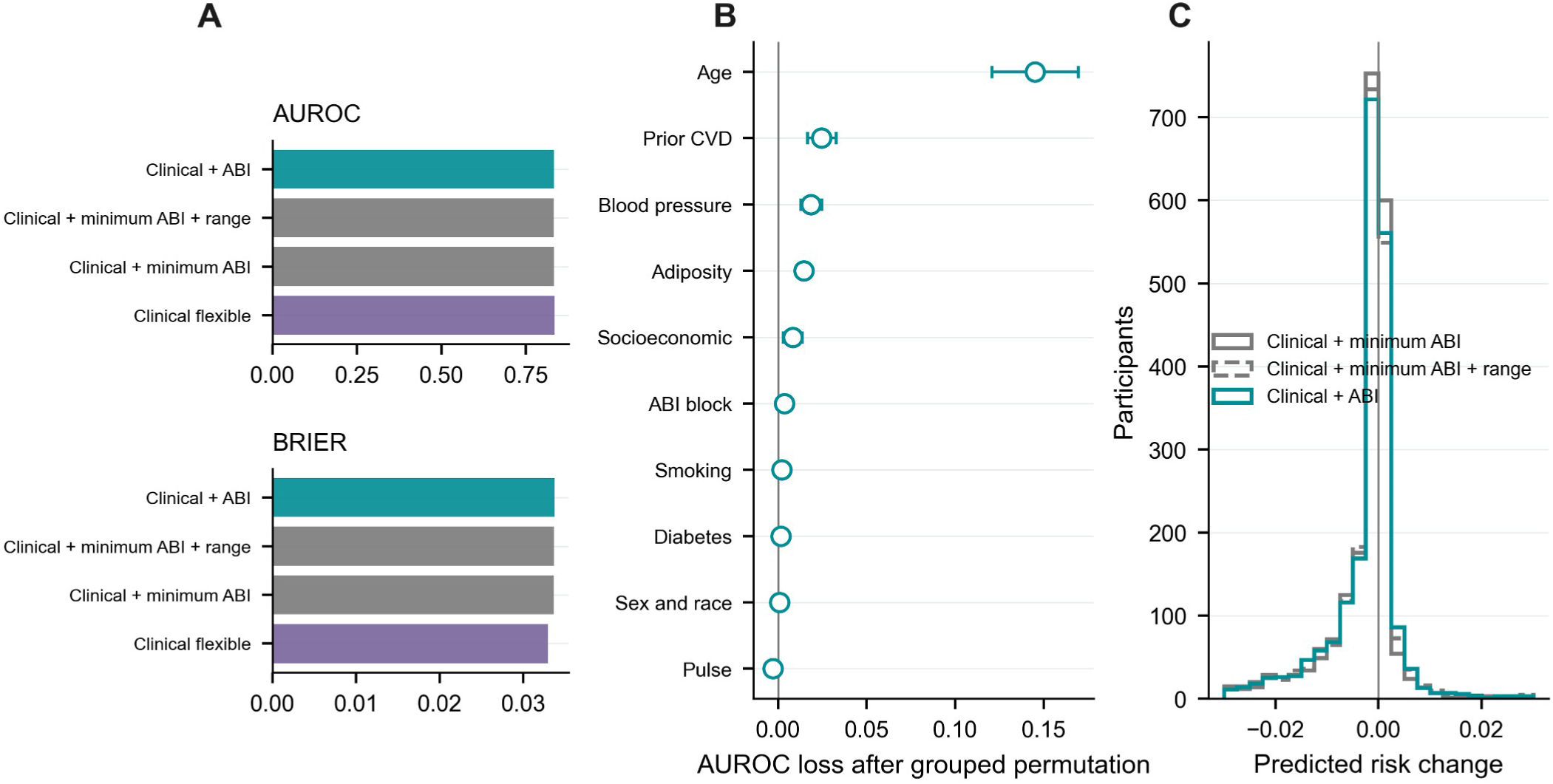
ABI ablation, grouped permutation, and individual prediction shifts. (A) Scores for the flexible clinical base, the same clinical covariates plus minimum ABI, the clinical base plus minimum ABI and ABI range, and the clinical + ABI model; lower Brier is better. (B) Mean AUROC loss after grouped permutation in the clinical + ABI model; error bars show one standard deviation across permutation repeats, not sampling confidence intervals. (C) Participant-level change in predicted 10-year CVD risk relative to the clinical flexible model.

**Figure 7.**
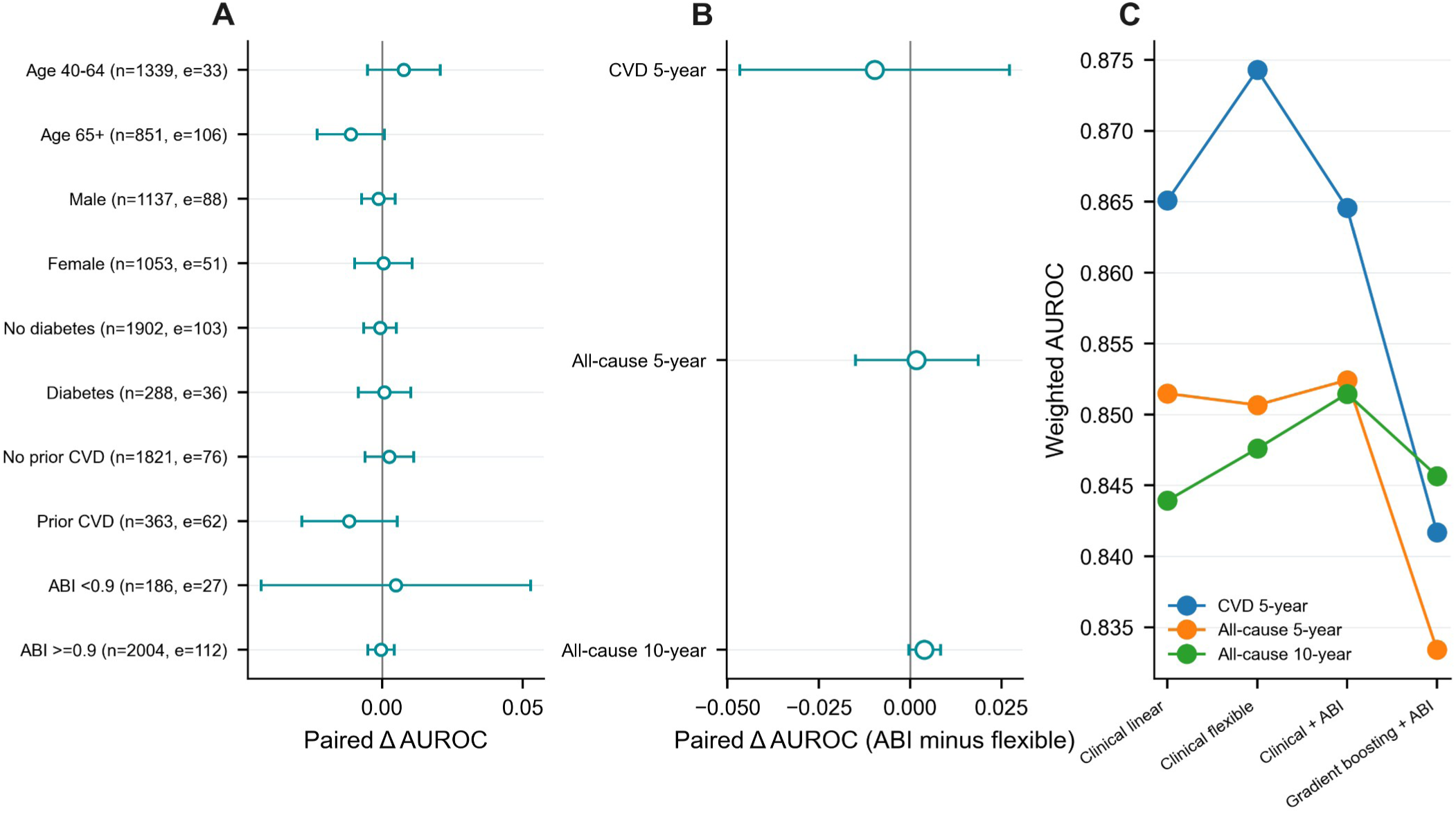
Exploratory subgroup and endpoint sensitivity analyses. (A) Paired AUROC difference (clinical + ABI minus clinical flexible) across subgroups; labels show subgroup sample sizes and events. (B) Paired difference for CVD 5-year, all-cause 5-year, and all-cause 10-year endpoints. (C) Weighted AUROC by model (clinical linear, clinical flexible, clinical + ABI, and gradient boosting + ABI) for CVD 5-year, all-cause 5-year, and all-cause 10-year endpoints; the endpoint names appear in the legend.

Two additional checks were added after internal review and are explicitly post hoc (Supplementary Table S1). In matched gradient-boosting models with and without ABI, ABI addition changed AUROC by 0.0097 (95% CI -0.0001 to 0.0194) and Brier score by -0.000216 (95% CI -0.000732 to 0.000301). Replacing the high-ABI feature threshold of ≥1.40 with >1.40 gave an ABI-versus-flexible-clinical AUROC difference of -0.0010 (95% CI -0.0046 to 0.0027). Neither check established improvement in both primary metrics. Figure 8 displays replicate-level uncertainty and the concentration of weighted target deaths across predicted-risk groups.

**Figure 8.**
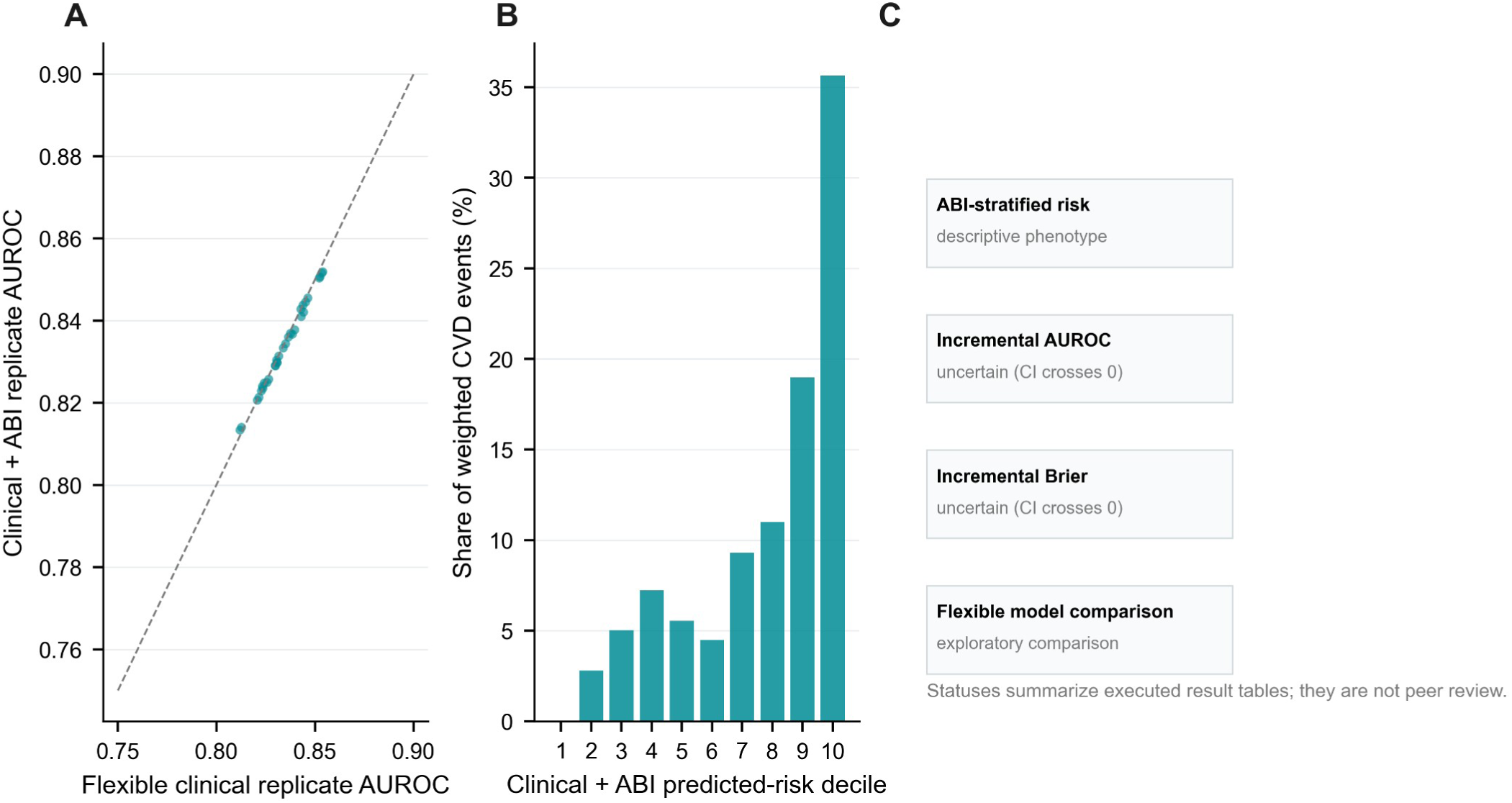
Replicate variability, weighted event concentration, and evidence summary. (A) Paired AUROC values across matched Fay balanced repeated replication (BRR) replicates; the diagonal denotes equality. (B) Percentage of total survey-weighted target deaths within ten approximately equal-count clinical + ABI predicted-risk deciles, numbered from 1 (lowest risk) to 10 (highest risk). (C) Summary of executed phenotype and paired-contrast results; labels describe these result tables and are not independent review judgments.

No formal fairness optimization or fairness assessment was performed beyond exploratory subgroup performance comparisons; these analyses do not establish equitable performance.

## Discussion

In this exploratory chronological evaluation, adding a bilateral ABI feature block to a flexible model of nonlaboratory clinical characteristics did not improve discrimination or Brier score for ten-year heart or cerebrovascular death. The paired AUROC difference was −0.0007 (conditional 95% CI −0.0047 to 0.0032), while the Brier difference was +0.0008 (−0.0003 to 0.0018). These findings concern specified fitted models and this ABI-observed population. They do not establish equivalence, negate ABI associations, or show that ABI lacks value for diagnosis, treatment selection or other endpoints.

The result narrows rather than overturns the prior literature. ABI association and incremental risk-stratification work already exists [5–11], and NHANES mortality prediction has also been studied using interpretable machine learning [12]. The practical contribution is an auditable matched comparison in which the unfavorable increment remains visible alongside calibration, proper scoring, selection diagnostics and design-aware paired intervals. The exploratory all-cause Brier signal should be tested in a new cohort rather than promoted as confirmation.

The investigators did not perform new outcome adjudication or predictor assessment. Original assessor demographics and blinding procedures are not available in the public analytic files; uniform secondary processing does not establish equivalent measurement quality across groups.

Several limitations constrain interpretation. The clinical comparator omits lipids, renal function, treatment details and validated comprehensive risk equations; it is a study benchmark rather than Framingham, PCE or PREVENT. One quarter of otherwise eligible participants lacked acceptable bilateral ABI, and observed ABI absence was associated with measured illness characteristics. Ordinary examination weights do not repair this measurement-selection process. The historical NHANES ABI protocol may differ from contemporary bilateral clinical protocols, the high-ABI tail is sparse, and public mortality causes/times may be perturbed. With only 270 development target deaths, model complexity and tuning instability are material concerns [18]; the small grid does not eliminate overfitting. Fixed-horizon competing deaths are non-events for the observed cumulative-risk estimand, not censored observations for a cause-specific hazard. Decision curves describe hypothetical threshold strategies without an intervention policy. Finally, the evaluation outcomes were inspected before this corrective revision; conditional BRR intervals do not include development uncertainty or analytical-choice uncertainty.

ABI summarizes limb pressure relationships. Experimental work on endothelial glycocalyx–lipoprotein interactions [23] and a review of biomechanical regulation of endothelial polarity [24] concern vascular dimensions that were not measured in this survey benchmark. Whether more direct endothelial or mechanobiological measurements add prognostic information is a future hypothesis; the present findings do not establish a mechanism for the observed ABI increment.

Translation to panvascular intervention would also require an explicit bridge between prognosis and treatment response. The proposed tri-ecological vessel–device framework [25] and FLOW panvascular-on-a-chip framework [26] offer conceptual and experimental directions for studying vascular and device interactions. Their proposed constructs were not measured here. A useful next study would pair externally validated risk prediction with appropriately measured biological or device-response endpoints; a population mortality score alone cannot establish which device or intervention benefits a patient.

## Conclusion

This reproducible public-data benchmark did not show a clear incremental prediction gain from bilateral ABI beyond a matched flexible clinical model for ten-year heart and cerebrovascular mortality. It supports further prospective and external evaluation of when ABI changes prediction, while providing no basis for changing screening, revascularization or other treatment decisions.

## Supporting information

Reproducible code, public-data manifests, aggregate results and reporting checklist

## Data Availability

Source data are publicly available from CDC/NCHS: NHANES 1999-2004 examination files and the 2019 public-use linked mortality files. Reconstruction code, source URLs and checksums, and aggregate analysis results are supplied in the supplementary reproducibility archive. Individual participant-level linked records and predictions are not uploaded to medRxiv; they can be reconstructed from the official public files. A persistent repository deposit is pending; no repository DOI has yet been issued.

https://wwwn.cdc.gov/nchs/nhanes/

https://ftp.cdc.gov/pub/Health_Statistics/NCHS/datalinkage/linked_mortality/

## Data availability

All input data are public NHANES examination files and the NCHS 2019 public-use linked mortality file. URLs, local source hashes, variable transformations, derived aggregate data and code are included in the reproducibility archive. Individual-row mortality-linked data are not redistributed beyond the CDC public-use terms.

## Code availability

The source archive includes build_data.py, describe.py, analyze.py, additional_checks.py, the numerical-figure scripts, manifests, aggregate result tables and run instructions. Participant-level predictions and saved fitted models are retained locally and can be recreated from official public inputs; they are not uploaded as medRxiv supplements. Results were regenerated after the corrected data build. A persistent repository deposit is pending; no repository DOI or reviewer URL has yet been issued.

## Author contributions

Lingsen You, Yujun Guo, Xinyu Zhong, Zisu Peng, Wentong Wang, Li Shen and Junbo Ge: Writing – review and editing. All authors reviewed and revised the AI-assisted text and approved the manuscript for posting. The computational assistance is described in the Methods and the declaration below.

## Ethics

This secondary analysis used only de-identified NHANES examination and public-use linked mortality data that were openly available before study initiation, without registration or application. The original NHANES 1999–2004 studies had NCHS ethics review under protocol 98-12 and participant consent [22]. No new participants were recruited or contacted, no identifiable records were accessed, and no attempt at re-identification was made. No new institutional ethics review was sought and no new approval or exemption decision was issued for this secondary analysis.

## DECLARATION OF GENERATIVE AI AND AI-ASSISTED TECHNOLOGIES

Codex assisted with LaTeX assembly, prose editing, and consistency checks. The authors reviewed and revised all AI-assisted text and take responsibility for the submitted manuscript.

---

**Table 1.** Analytic cohort characteristics.

| Characteristic | Development n = 4,617 | Evaluation n = 2,190 |
|---|---|---|
| Age, years | 55.75 (11.82) | 55.73 (11.62) |
| BMI, kg/m² | 28.15 (5.76) | 28.30 (5.51) |
| Systolic BP, mmHg | 127.99 (19.71) | 127.90 (18.86) |
| Male | 2,370 / 4,617; 48.3% | 1,137 / 2,190; 49.6% |
| Female | 2,247 / 4,617; 51.7% | 1,053 / 2,190; 50.4% |
| Never smoker | 2,127 / 4,609; 46.3% | 984 / 2,189; 45.1% |
| Former smoker | 1,558 / 4,609; 32.8% | 760 / 2,189; 33.6% |
| Current smoker | 924 / 4,609; 20.8% | 445 / 2,189; 21.3% |
| Reported diagnosed diabetes | 543 / 4,615; 8.5% | 288 / 2,190; 9.8% |
| Reported prior cardiovascular disease | 633 / 4,583; 11.5% | 363 / 2,184; 12.8% |
| Minimum ABI | 1.11 (0.12) | 1.09 (0.13) |
| Heart/cerebrovascular death, 10 years | 270 / 4,617; 3.9% | 139 / 2,190; 3.8% |
| All-cause death, 10 years | 863 / 4,617; 13.9% | 415 / 2,190; 11.8% |

Continuous values are survey-weighted means (SD). Categorical cells give raw count/observed denominator and weighted percentage; these are distinct quantities. Missing values are excluded from the variable-specific denominator. Full descriptive and missingness tables accompany the code.

**Table 2.** Primary evaluation performance.

| **Model** | **AUROC** | **Average precision** | **Brier** | **CITL** | **Slope** |
|---|---|---|---|---|---|
| Clinical linear | 0.830<br>(0.772, 0.888) | 0.196<br>(0.139, 0.253) | 0.0331<br>(0.0259, 0.0403) | -0.037<br>(-0.275, 0.200) | 0.904<br>(0.696, 1.112) |
| Clinical flexible | 0.834<br>(0.786, 0.882) | 0.203<br>(0.150, 0.255) | 0.0329<br>(0.0256, 0.0402) | -0.036<br>(-0.278, 0.206) | 0.895<br>(0.716, 1.074) |
| Clinical + ABI | 0.834<br>(0.788, 0.879) | 0.208<br>(0.137, 0.279) | 0.0337<br>(0.0269, 0.0405) | -0.058<br>(-0.299, 0.184) | 0.817<br>(0.662, 0.972) |
| Boosting + ABI | 0.822<br>(0.770, 0.874) | 0.207<br>(0.123, 0.291) | 0.0331<br>(0.0264, 0.0397) | -0.002<br>(-0.235, 0.232) | 0.822<br>(0.658, 0.985) |

95% intervals use 32 Fay BRR replicates, rho = 0.5 and t(15), conditional on the fitted models. CITL is calibration-in-the-large with prediction-logit slope fixed to one; slope is jointly estimated with an intercept. Lower Brier is better.

