## Supplementary methods, Tables S1-S4 and Figures S1-S6 for "Incremental value of bilateral ankle-brachial index for ten-year heart and cerebrovascular mortality prediction: an exploratory temporal benchmark in NHANES"

31 October 2026

Incremental value of bilateral ankle-brachial index for ten-year heart and cerebrovascular mortality prediction: an exploratory temporal benchmark in NHANES

### S1. Corrective data reconstruction

The original draft is superseded. The corrected build retains actual age 77 and waist 77/99 measurements, uses questionnaire-specific missing codes, respects never-smoker skips, preserves uncertain comorbidity responses as missing, and corrects pooled examination weights. One death with unavailable underlying cause within ten years was excluded. The source manifest records 27 public inputs with URLs, byte counts and SHA-256 hashes. Results/data_audit.json and results/flow.csv record exact counts. No numerical result in this revision comes from synthetic or simulated participants.

The initial working plan emphasized five-year all-cause survival, while the earlier executed draft used ten-year heart/cerebrovascular death. The corrective protocol retained the latter displayed endpoint after those outcomes had already been examined. Accordingly, all present analyses are exploratory. A dated additional-check amendment records the subsequent matched boosting and high-ABI boundary checks.

### S2. Variables and missing information

Table S4 provides the source fields and transformations. The analytic cohort requires observed bilateral ABI, but does not require complete clinical predictors. Continuous clinical values are imputed using development medians and accompanied by missingness indicators. Nominal variables use a distinct missing category and one-hot encoding. The same fitted transformations are applied unchanged to evaluation data. Complete-case sensitivity excludes any missing clinical predictor in either period and refits all model classes. The included-versus-excluded comparison is descriptive and does not correct for selection into observed ABI.

The diabetes flag treats borderline diabetes as not meeting the reported diabetes definition. Prior cardiovascular disease includes reported congestive heart failure, coronary heart disease, angina, myocardial infarction or stroke. These self-reported measures differ from adjudicated clinical diagnoses. Blood-pressure predictors are analyst-defined means of positive available readings, including the first, rather than the CDC reported-average algorithm. These choices limit direct comparison with established clinical scores.

**S3. Model selection and leakage checks**

Regression uses C in {0.01, 0.1, 1, 10}. Flexible regression applies cubic splines with four development-quantile knots to age, systolic pressure and body mass index, plus minimum ABI when included. Boosting uses 150 iterations, learning rate 0.04, minimum leaf size 35, no early stopping, and a four-combination grid of 7/15 leaves and L2 of 1/10. Hyperparameters minimize weighted Brier score on 2001–2002 after fitting 1999–2000, then all development participants are used for refitting. Training weights are normalized to mean one. Development-grid scores and selected settings are recorded in results/tuning_results.csv and results/selected_hyperparameters.json; the additional checks use results/additional_tuning.csv and results/additional_manifest.json. Endpoint and missingness sensitivities repeat development-only tuning; evaluation outcomes do not enter preprocessing or selection.

Model reload checks compare predictions to retained tables using a numerical tolerance and recalculate performance from outcomes, predictions and weights. Such checks can detect implementation inconsistency but cannot remove bias from previously inspected test outcomes, guarantee clinical validity, or establish independent replication by human investigators. Follow-up, causes of death, mortality status, participant IDs, cycle labels, survey weights and survey design identifiers are excluded from predictors.

#### S4. Survey variance and paired comparisons

Let theta denote a full-weight estimate and theta_r the estimate from replicate r. With R=32 and Fay rho=0.5, variance is sum_r (theta_r − theta)^2 / [R(1−rho)^2]. Intervals are theta ± t(0.975,15) times the square root of that variance. Pairwise differences are recomputed inside each common replicate before applying the same formula; marginal confidence limits are never subtracted. This is conditional evaluation-sampling uncertainty, without model-development, tuning or linkage uncertainty.

The evaluation cycle contains 15 strata, each with two masked PSUs. A 32-order Hadamard matrix supplies columns 1 through 14 and 16 using zero-based indexing, with −1/+1 coding for the two PSUs. All 32 patterns are unique. The alternative first-15-column selection had only 16 unique patterns and was replaced before the final calculations. This design correction was based on pattern uniqueness, not a preferred result. Domain estimates retain the full design map and subset only at metric evaluation. Weight scaling leaves normalized metric point estimates unchanged.

Calibration-in-the-large estimates an intercept with prediction-logit slope fixed to one. Calibration slope is estimated jointly with an intercept. Fixed-bin calibration uses weighted observed and predicted risks; bins with fewer than five events have observed-risk estimates and intervals suppressed. AUROC uses weighted concordance; average precision is the weighted stepwise precision–recall summary, not trapezoidal area.

Decision curves use weighted event and non-event proportions with the threshold odds penalty; no treatment effect is estimated.

### S5. Exploratory extensions

Endpoint analyses examine five-year heart/cerebrovascular mortality and five-/ten-year all-cause mortality. Complete-case and unweighted refits, ABI-block ablations, subgroup contrasts, selection comparisons, missingness, weights, calibration, decision curves, and grouped ABI permutation are retained. Permuting the entire ABI block with a common row permutation preserves its internal deterministic relationships. Repeated permutation spread is a diagnostic of predictive reliance, not a population confidence interval or a causal effect.

Post hoc checks compare matched boosting models with and without ABI and change the either-leg high-ABI definition from ≥1.40 to >1.40. Primary outputs remain unchanged. Minimum ABI exactly 0.90 occurs in 28 development and 14 evaluation participants; maximum ABI exactly 1.40 occurs in eight and one respectively. The minimum-ABI ≥1.40 evaluation group contains only six participants and two target events, so its risk estimate and interval are suppressed. No interaction tests or multiplicity-controlled subgroup discoveries are claimed.

### S6. Reporting and reproducibility

Every quantitative panel is generated by code from saved computed tables. The source archive contains reconstruction and analysis scripts, aggregate results, reference and source manifests, and execution instructions. It excludes participant-level linked records and predictions from the medRxiv upload. These can be regenerated locally from official CDC inputs. The figure design was informed by public Ling Yang research figures, but no images, mechanisms or numerical findings were copied. Internal agent checks cover data provenance, methods, leakage, references, scientific claims, figures, layout and reproducibility; they are internal verification, not external peer review.

This analysis uses the same public NHANES cycles as a separate author-group project predicting low ABI from clinical variables. Here ABI is a predictor and mortality is the outcome. The companion project's current submission status must be disclosed accurately when known. Another Codex conversation contains an earlier revision of this same mortality study; it is a version of this project, not a separate study eligible for a second submission.

### S7. Ethics and author review

Original NHANES 1999–2004 oversight used NCHS protocol 98-12 and participant consent. This secondary analysis uses only de-identified examination and public-use linked mortality data openly available before study initiation, without registration or application. No new participants were recruited or contacted, no identifiable records were accessed, and no attempt at re-identification was made. No new institutional ethics review was sought and no new approval or exemption decision was issued for this analysis. All seven authors participated in manuscript review and editing, reviewed and revised the AI-assisted text, and approved the manuscript for posting. No repository DOI or reviewer URL has been issued for this package.

### DECLARATION OF GENERATIVE AI AND AI-ASSISTED TECHNOLOGIES

Codex assisted with LaTeX assembly, prose editing, and consistency checks. The authors reviewed and revised all AI-assisted text and take responsibility for the submitted manuscript.

### Additional post hoc checks

#### Table S1. Matched boosting and ABI boundary sensitivity

| Contrast | Metric | Difference | 95% CI |
|---|---|---|---|
| Boosting + ABI minus Boosting clinical | AUROC | 0.009678 | -0.000063 to 0.019420 |
| Boosting + ABI minus Boosting clinical | Average precision | 0.015218 | -0.019215 to 0.049651 |
| Boosting + ABI minus Boosting clinical | Brier score | -0.000216 | -0.000732 to 0.000301 |
| Strict-high ABI minus Clinical flexible | AUROC | -0.000957 | -0.004647 to 0.002732 |
| Strict-high ABI minus Clinical flexible | Average precision | 0.005584 | -0.028270 to 0.039438 |
| Strict-high ABI minus Clinical flexible | Brier score | 0.000783 | -0.000269 to 0.001836 |
| Strict-high ABI minus Clinical + ABI | AUROC | -0.000213 | -0.000776 to 0.000351 |
| Strict-high ABI minus Clinical + ABI | Average precision | 0.000019 | -0.000624 to 0.000661 |
| Strict-high ABI minus Clinical + ABI | Brier score | -0.000001 | -0.000008 to 0.000005 |

Clinical + ABI denotes the flexible clinical model extended with the ABI block. Strict-high ABI uses that model with either-leg high ABI defined as >1.40 instead of ≥1.40. Boosting clinical omits ABI from the matched gradient-boosting model. Differences follow the stated first model minus second model; positive AUROC or average-precision differences and negative Brier-score differences indicate improvement. Intervals are conditional 95% Fay-BRR intervals.

#### Table S2. Exploratory endpoint and missing-data contrasts

| Analysis | Metric | Difference | 95% CI |
|---|---|---|---|
| 5-year heart/cerebrovascular death | AUROC | -0.00973 | -0.04663 to 0.02716 |

| Analysis | Metric | Difference | 95% CI |
|---|---|---|---|
| 5-year heart/cerebrovascular death | Brier score | 0.00001 | -0.00039 to 0.00040 |
| 5-year all-cause death | AUROC | 0.00175 | -0.01503 to 0.01854 |
| 5-year all-cause death | Brier score | 0.00017 | -0.00084 to 0.00119 |
| 10-year all-cause death | AUROC | 0.00386 | -0.00050 to 0.00823 |
| 10-year all-cause death | Brier score | -0.00110 | -0.00197 to -0.00023 |
| Unweighted refit | AUROC | -0.00130 | -0.02065 to 0.01804 |
| Unweighted refit | Brier score | 0.00038 | -0.00061 to 0.00137 |
| Complete-case refit | AUROC | -0.00263 | -0.00965 to 0.00440 |
| Complete-case refit | Brier score | 0.00090 | -0.00019 to 0.00200 |
| Complete cases, original models | AUROC | -0.00165 | -0.00767 to 0.00436 |
| Complete cases, original models | Brier score | 0.00103 | -0.00012 to 0.00219 |

Every difference is Clinical + ABI minus Clinical flexible. Weighting and complete-case analyses retain the primary 10-year heart/cerebrovascular-death endpoint; alternate endpoints are named in the rows. Positive AUROC differences and negative Brier-score differences indicate improvement. Intervals are conditional 95% Fay-BRR intervals; all comparisons are exploratory.

### Table S3. ABI block ablation contrasts

| Contrast | Metric | Difference | 95% CI |
|---|---|---|---|
| Clinical + ABI minus Clinical flexible | AUROC | -0.00074 | -0.00469 to 0.00320 |
| Clinical + ABI minus Clinical flexible | Brier score | 0.00078 | -0.00027 to 0.00184 |
| Clinical + minimum ABI minus Clinical flexible | AUROC | -0.00056 | -0.00407 to 0.00294 |
| Clinical + minimum ABI minus Clinical flexible | Brier score | 0.00070 | -0.00035 to 0.00175 |
| Clinical + minimum ABI and range minus Clinical + minimum ABI | AUROC | -0.00125 | -0.00373 to 0.00122 |
| Clinical + minimum ABI and range minus Clinical + minimum ABI | Brier score | 0.00007 | -0.00008 to 0.00022 |
| Clinical + ABI minus Clinical + minimum ABI and range | AUROC | 0.00107 | -0.00238 to 0.00453 |
| Clinical + ABI minus Clinical + minimum ABI and range | Brier score | 0.00002 | -0.00006 to 0.00010 |

The minimum-ABI and minimum-plus-range models extend the flexible clinical model. Clinical + ABI additionally includes the either-leg high-ABI indicator. Differences follow the stated first model minus second model. Positive AUROC differences and negative Brier-score differences indicate improvement. Intervals are conditional 95% Fay-BRR intervals; all comparisons are exploratory.

### Table S4. Public variables and transformations

| Derived variable | Public source | Definition |
|---|---|---|
| age | RIDAGEYR | Years as released; 77 is a valid measurement; public top-coding is retained. |
| sex | RIAGENDR | 1 male; 2 female; other values missing. |
| race | RIDRETH1 | Nominal categories 1–5; no ordinal numeric effect. |
| education | DMDEDUC2 | Nominal categories 1–5; refusal/unknown values missing. |
| poverty | INDFMPIR | Public poverty-income ratio, as measured. |
| bmi; waist | BMXBMI; BMXWAIST | Continuous measurements; waist 77/99 cm retained. |
| sbp; dbp | BPXSY1–4; BPXDI1–4 | Arithmetic mean of all positive available readings, including reading 1. Not the CDC reported-average algorithm. |
| pulse | BPXPLS | Continuous recorded pulse rate. |
| diabetes | DIQ010 | 1 yes → 1; 2 no or 3 borderline → 0; other responses missing. |
| hypertension | BPQ020 | 1 yes → 1; 2 no → 0; other responses missing. |
| prior_cvd | MCQ160B–F | Any yes → 1; all five explicitly no → 0; otherwise missing. |
| smoking | SMQ020; SMQ040 | Never: SMQ020=2; former: SMQ020=1 and SMQ040=3; current: SMQ020=1 and SMQ040=1/2; otherwise missing. |

| Derived variable | Public source | Definition |
|---|---|---|
| abi_min; abi_max | LEXLABPI; LEXRABPI | Minimum/maximum after both legs observed in [0.20, 2.50]. No unilateral substitution. |
| abi_range | Derived ABI | Absolute left-right difference, equivalent to maximum minus minimum. |
| abi_high_any | Derived ABI | Either leg ≥1.40; the additional sensitivity uses >1.40. |
| abi_low | Derived ABI | Minimum ABI <0.90; descriptive grouping only, not a primary predictor. |
| survey_weight | WTMEC4YR; WTMEC2YR | 1999–2002: 2/3 × WTMEC4YR; 2003–2004: 1/3 × WTMEC2YR. |
| design fields | SDMVSTRA; SDMVPSU | Used for survey variance only; excluded from predictors. |
| cvd_10y; cvd_5y | MORTSTAT; UCOD; PERMTH_EXM | Death from code 001/005 by 120/60 months. Other-cause deaths are non-events. Unknown cause within horizon excluded. |
| all_10y; all_5y | MORTSTAT; PERMTH_EXM | Any death by 120/60 months. Living participants require follow-up through the horizon. |
| SEQN; cycle; split | DEMO and file names | Joining, chronological allocation and audit only; never predictors. |

### Supplementary Figure S1

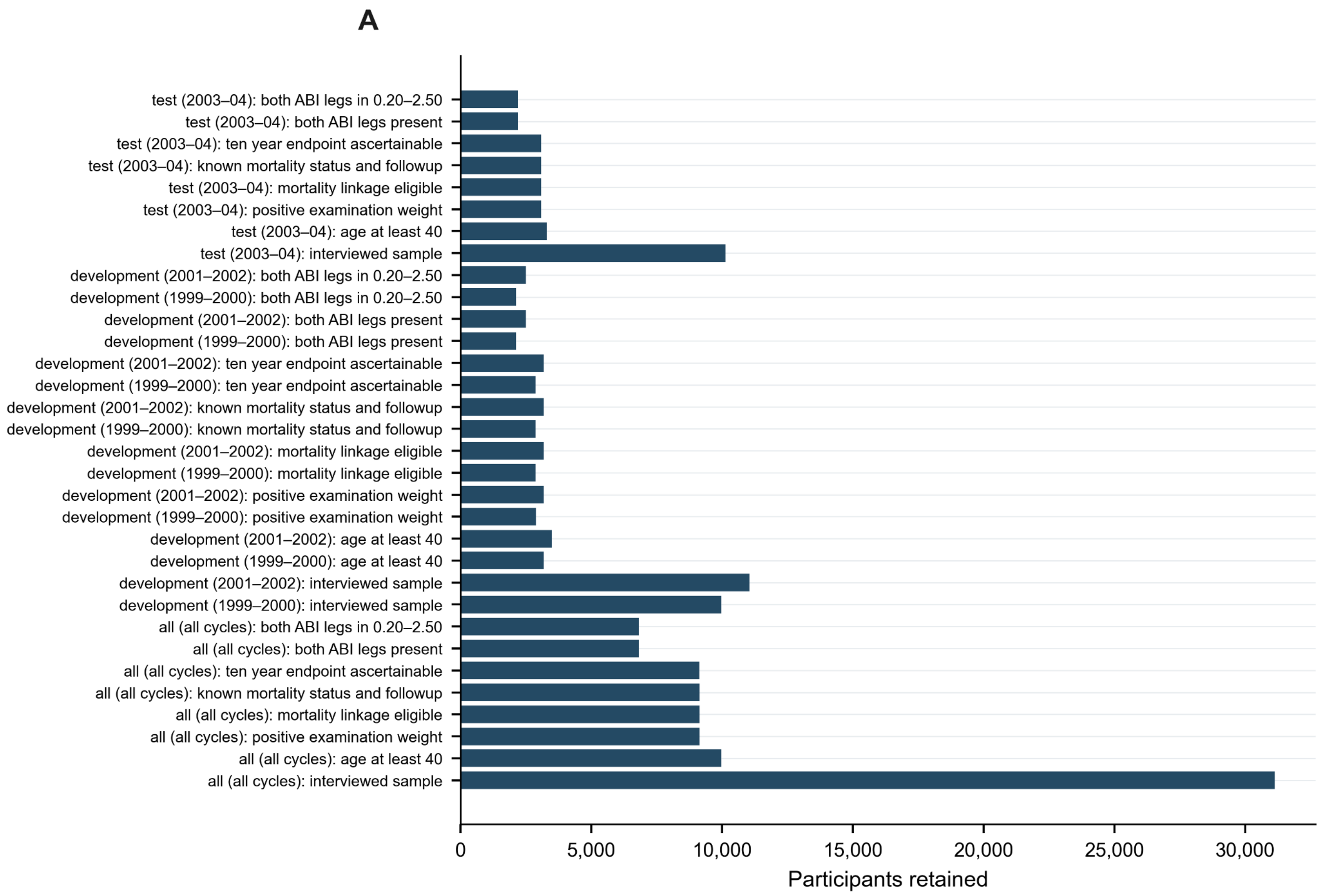


Supplementary Figure S1. Sequential participant counts retained across source filtering for each split and survey cycle. Development cycles are shown separately, alongside the combined all-cycle cohort and chronological test cycle.

**Supplementary Figure S2**

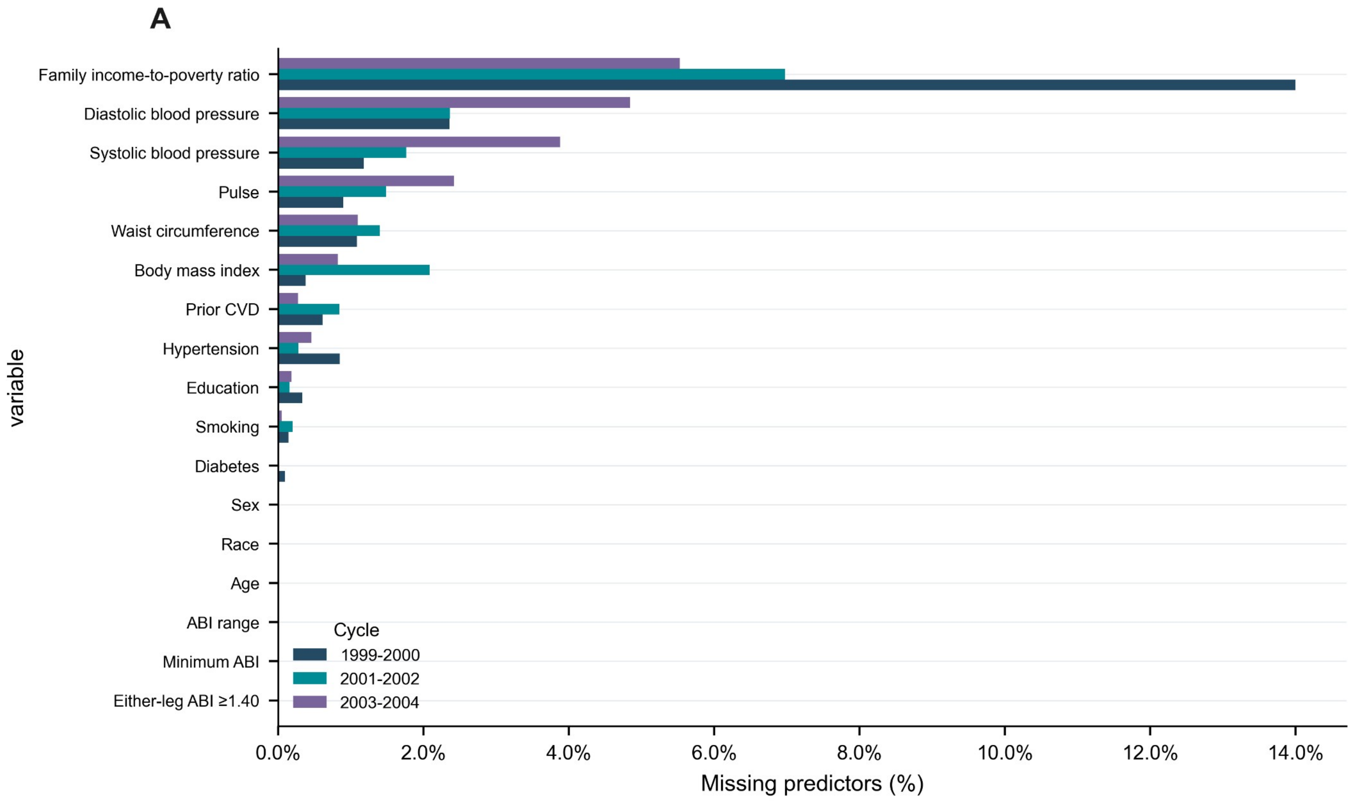


Supplementary Figure S2. Percentage of missing values for each predictor by survey cycle. Bars are derived from the saved missingness table; cycle colors are labeled in the legend.

**Supplementary Figure S3**

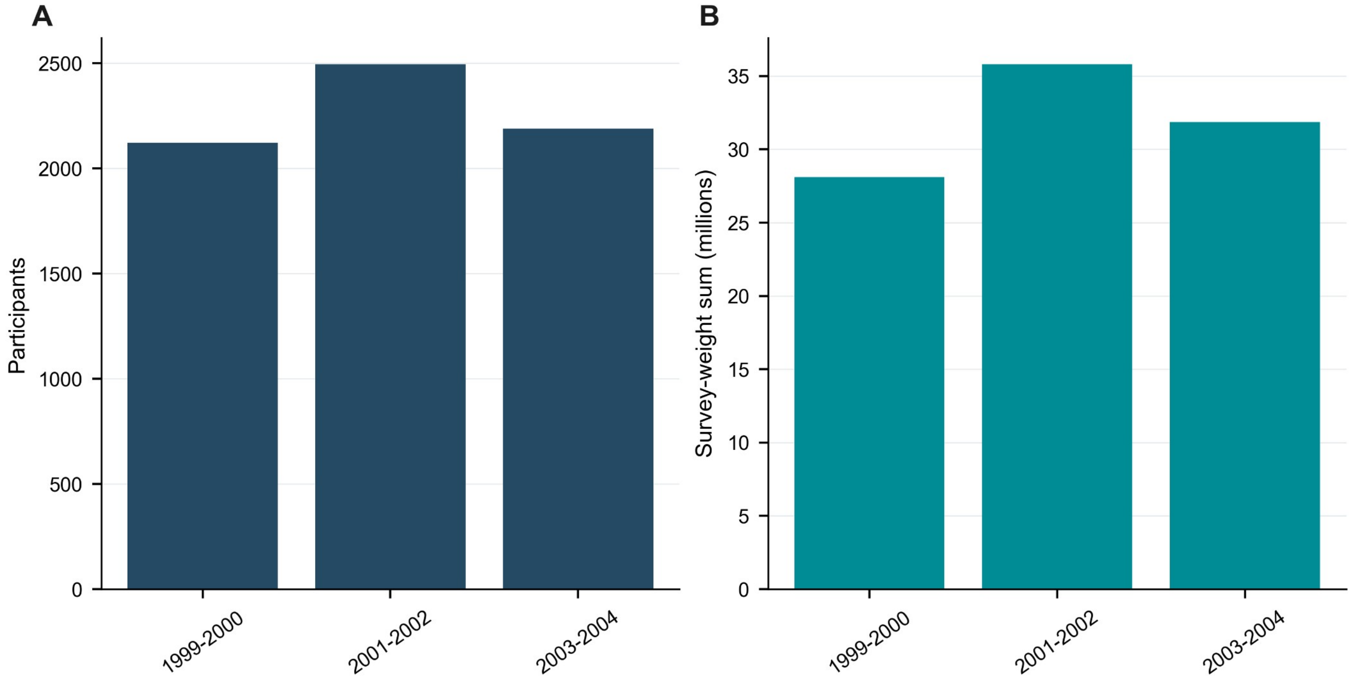


Supplementary Figure S3. (A) Analytic participant counts by survey cycle. (B) Sum of examination survey weights by cycle, in millions.

### Supplementary Figure S4

A

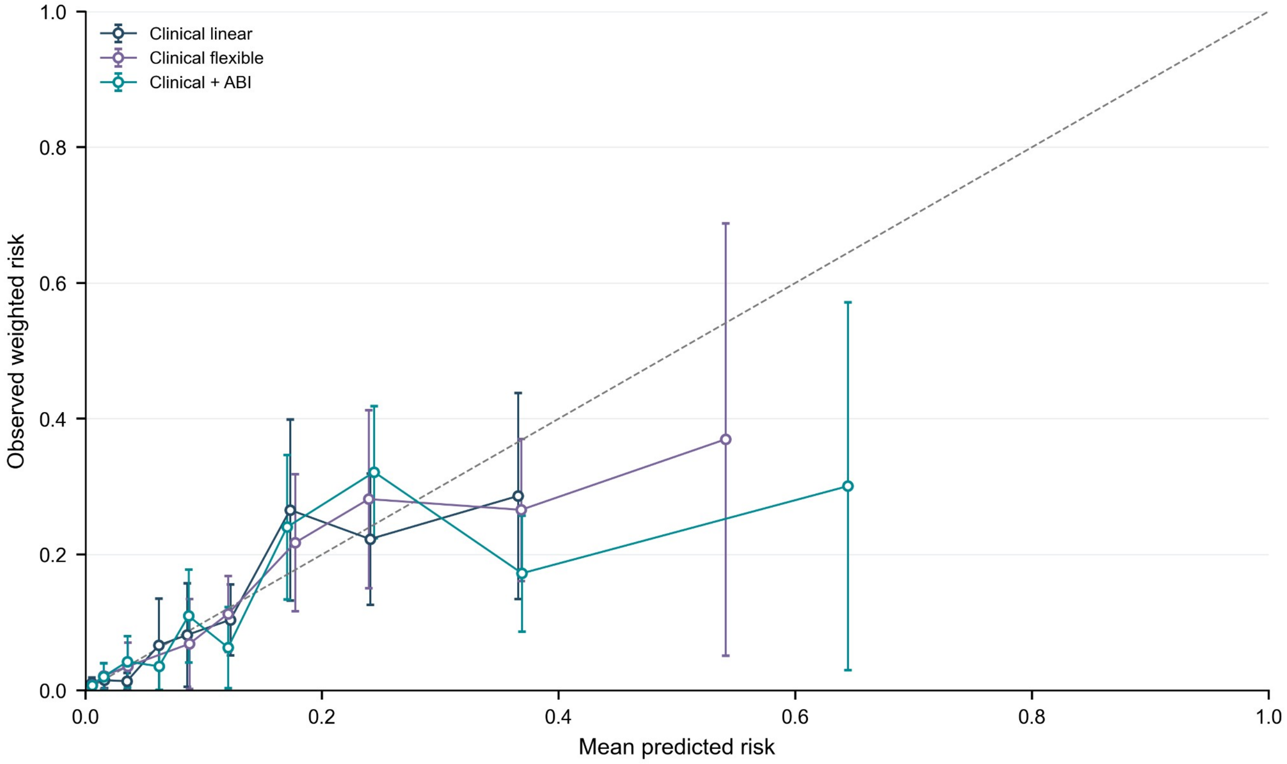


Supplementary Figure S4. Full-scale calibration across all supported risk bins for the clinical linear, clinical flexible, and clinical + ABI models. Points and intervals show observed weighted risk and its saved interval; the dashed diagonal is perfect calibration.

### Supplementary Figure S5

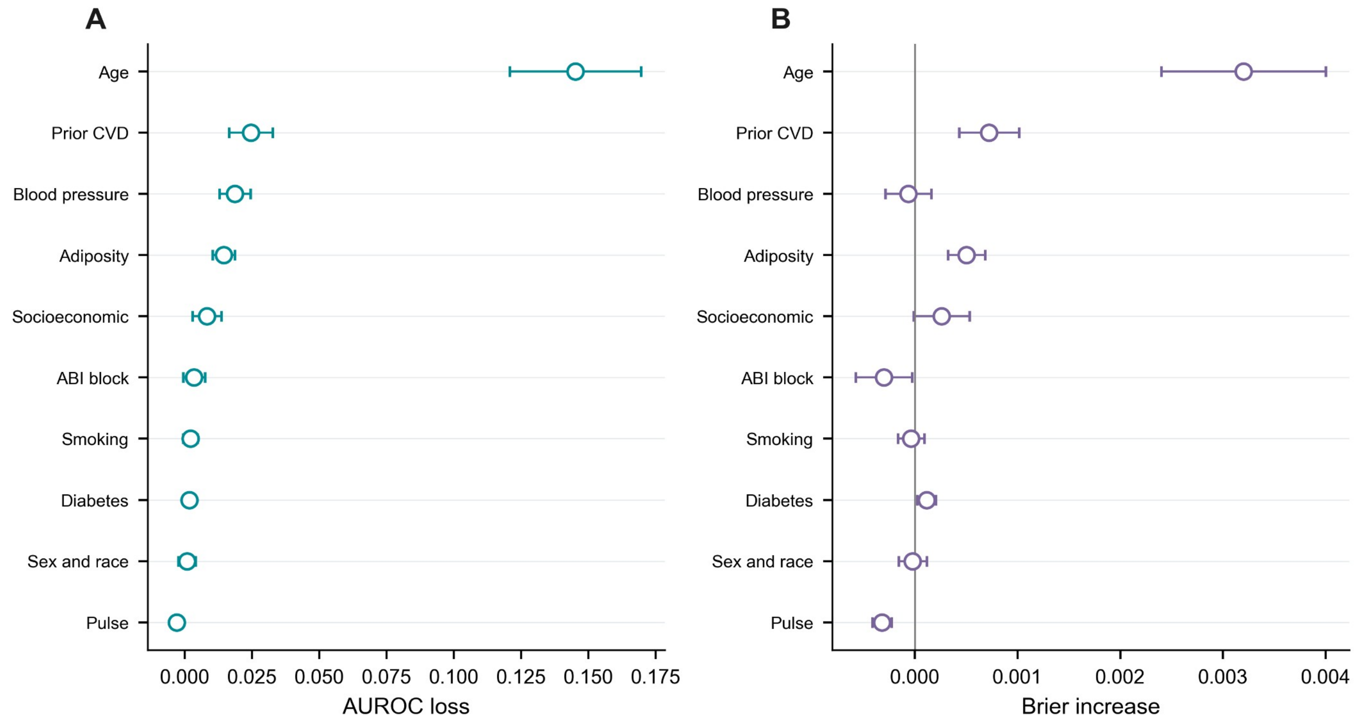


Supplementary Figure S5. Repeat-level grouped permutation results for the clinical + ABI model only. (A) AUROC loss after permuting each predictor group. (B) Brier-score increase. Horizontal intervals show repeat-to-repeat standard deviations and describe permutation variability, not sampling uncertainty or causal effects.

### Supplementary Figure S6

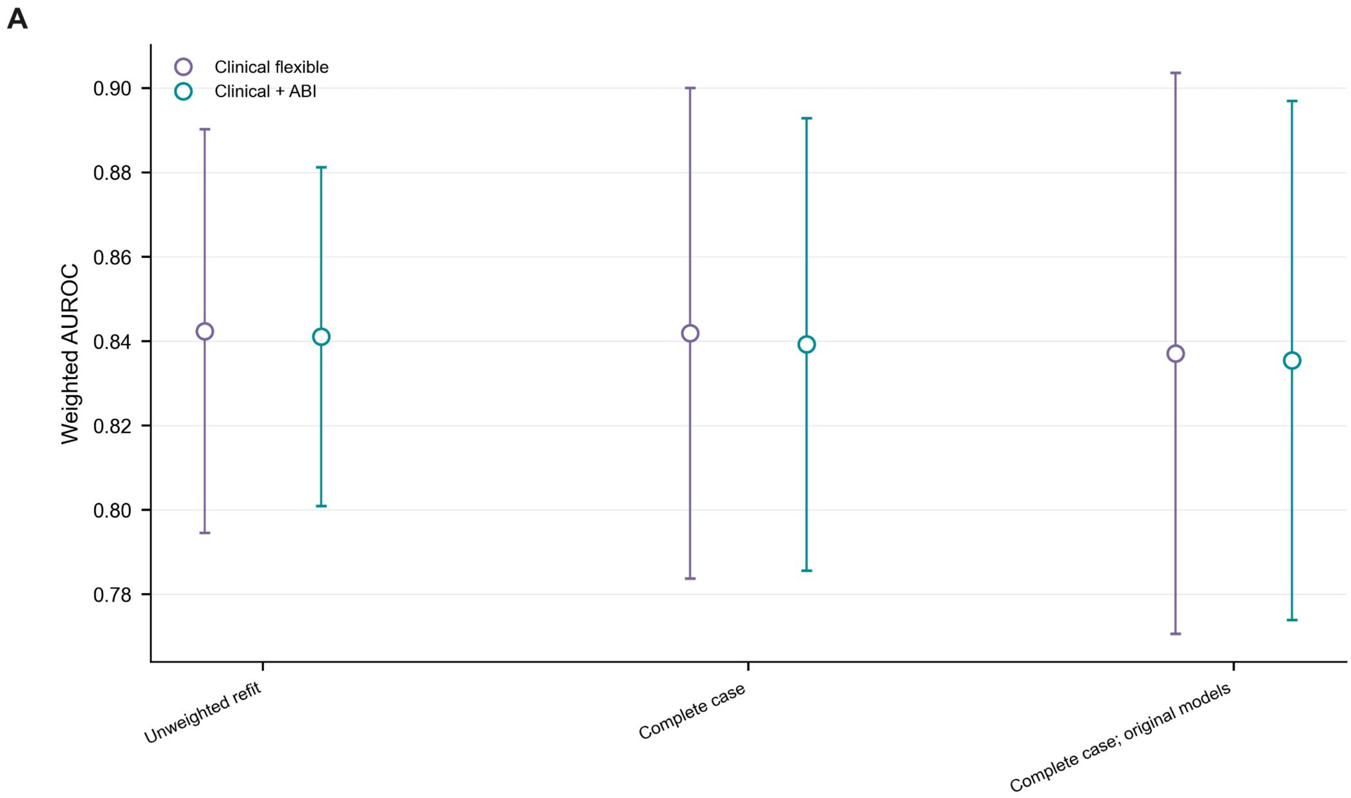


Supplementary Figure S6. Weighted AUROC for clinical flexible and clinical + ABI models under unweighted refitting and complete-case sensitivity analyses. Points and intervals are reported in the saved sensitivity metric table.
